# Prediction of Covid-19 spreading and optimal coordination of counter-measures: From microscopic to macroscopic models to Pareto fronts

**DOI:** 10.1101/2020.12.01.20241885

**Authors:** Hanna Wulkow, Tim Conrad, Nataša Djurdjevac Conrad, Sebastian A. Mueller, Kai Nagel, Christof Schuette

**Affiliations:** Zuse Institute Berlin, 14195, Germany; Department of Mathematics and Computer Science, Freie Universität Berlin, 14195, Germany; Transport Systems Planning and Transport Telematics, TU Berlin, Berlin, Germany

## Abstract

The Covid-19 disease has caused a world-wide pandemic with more than 60 million positive cases and more than 1.4 million deaths by the end of November 2020. As long as effective medical treatment and vaccination are not available, non-pharmaceutical interventions such as social distancing, self-isolation and quarantine as well as far-reaching shutdowns of economic activity and public life are the only available strategies to prevent the virus from spreading. These interventions must meet conflicting requirements where some objectives, like the minimization of disease-related deaths or the impact on health systems, demand for stronger counter-measures, while others, such as social and economic costs, call for weaker counter-measures. Therefore, finding the optimal compromise of counter-measures requires the solution of a multi-objective optimization problem that is based on accurate prediction of future infection spreading for all combinations of counter-measures under consideration. We present a strategy for construction and solution of such a multi-objective optimization problem with real-world applicability. The strategy is based on a micro-model allowing for accurate prediction via a realistic combination of person-centric data-driven human mobility and behavior, stochastic infection models and disease progression models including micro-level inclusion of governmental intervention strategies. For this micro-model, a surrogate macro-model is constructed and validated that is much less computationally expensive and can therefore be used in the core of a numerical solver for the multi-objective optimization problem. The resulting set of optimal compromises between counter-measures (Pareto front) is discussed and its meaning for policy decisions is outlined.

## 1 Introduction

The Covid-19 disease, caused by infection with the virus SARS-CoV-2, was first detected end of 2019 in China before spreading all over the world within a few months. Symptoms of an infection include fever, dry cough and fatigue, as well as sore throat and loss of taste and smell [7]. While the majority of patients are either asymptomatic or have mild symptoms, for around 5.7 % (data from the US [4]) of people the disease can lead to life-threatening conditions such as pneumonia which in many cases leads to hospitalization. Out of these, about 40 % are moved to the intensive care unit (ICU). Current studies show that approximately 3 % of people die from a Covid-19 infection worldwide [2, 38]. This so-called mortality rate is higher for people with underlying health conditions, such as COPD, obesity, type 2 diabetes mellitus or serious heart conditions [5] and lower for children and young adults [36].

Compared to the average mortality (death per 100,000 population) of influenza, the average mortality of Covid-19 is significantly higher (2.0 for influenza vs. 62.5 for Covid-19 e.g. in the US [6]). This, together with the high infectiousness of the SARS-CoV-2 virus has led to national restrictions of the public life in order to prevent the health care system from becoming overwhelmed. These restrictions include the closure of schools and universities, shops and restaurants, social distancing including wearing of masks, and so-called national “lockdowns”, where people being prohibited from leaving their home except for work and essential shopping. Further, during a lockdown, all non-essential shops are closed and public gatherings of people not from the same household are forbidden [1].

While lockdowns in particular have had a strong reduction effect on the infection numbers, they come with severe social and economic repercussions. Stopping people from leaving their homes and interacting with others face-to-face can lead to a decline in physical and mental health, while whole industries such as the hospitality industry have recorded extreme losses in the months since March 2020 [9]. Since the first wave of lockdowns and severe restrictions in March, April and May, governments have been trying to find a way to keep infection numbers small while keeping the economy strong and enabling people to live in a quasi-normal way. This has proven to be a difficult task, mainly since it involves different conflicting objectives, e.g., keep the public acceptance for the restrictions high, and stop the economy from stalling, while slowing the spreading and preventing the health care systems from becoming overwhelmed.

It would be useful to have a way of computing which combination of government restrictions and policies (from here on called “counter-measures”) form *optimal* compromises between the conflicting objectives. To do this, the relationship between counter-measures and their effect on the infection behavior has to be predicted, in particular for combinations of counter-measures that have not been tried in reality yet or are hard to implement. Because in most countries several measures were implemented at once, it is difficult to guess their individual effects. Based on accurate prediction one would be able to approximate the Pareto set [23] of Pareto optimal points with respect to the conflicting objectives. While being possible in principle, such an endeavor would require accurate *and* easily computable prediction schemes for all possible combinations of counter-measures. Based on these, one could approximate the Pareto set via available techniques for multi-objective optimization [8].

For accurate prediction of counter-measures, in particular also for combinations that have never been tried in reality, so-called agent-based models have been proposed that permit modelling the infection spreading on the basis of the spatio-temporal interaction of individuals (“agents”) with realistic mobility and behavioral patterns. Several such ABM have been described in the literature, recently specifically for the spread of Covid-19, e.g., [24] and [15].

In this paper, we report on an agent-based mobility model for Berlin, that has been developed based on recent publications on the mechanism of Covid-19 spreading, and allows to simulate the dynamics of the infection spreading in the city of Berlin on a micro-level (see [24]). With this model (called the “ABM” from here on) the infection numbers of, e.g., exposed, contagious, symptomatic and hospitalized patients, can be predicted for all measures individually and accurately. However, the ABM is computationally very expensive since in every single time step the interaction of several 100.000 agents has to be computed. This very high computational effort prevents the use of the ABM as the prediction core of a multi-objective optimization scheme.

Therefore, we have developed an accompanying macro-model based on differential equations (called the “ODE model” subsequently) that uses the ABM results to approximate the change in the infection behavior for different counter-measures. Computationally, the ODE model is computationally cheap and can thus serve as the prediction core for computing the Pareto sets wrt. a set of conflicting objectives.

We will discuss below, whether the prediction quality of the ODE model is sufficient for this task. To this end, we will first demonstrate that the ODE model is able to reproduce the data coming from real-world observations of the Covid-19 pandemics, as well as for simulation data originating from the ABM, after appropriate parameter estimation. The ODE model contains 10 kinetic parameters, some of them being related to the basic properties of Covid-19 (like the typical time of showing symptoms or the hospitalization rate) and others depending on the counter-measures in effect (like the average infection rate). We observe that only the latter parameters do change if the ODE model is fitted to data simulating different counter-measure combinations.

Next, we approximate the change of these parameters if the underlying combinations of counter-measures are changed. That is, we consider the ODE parameters as functions on the counter-measure space (that is, the abstract space of all possible counter-measure combinations). By learning this function from data, the ODE model is turned into a “stand-alone” model that can be run for all possible combinations of counter-measures without the requirement to fit it to appropriate data first. This then allows for the predictive use of the stand-alone ODE model for different counter-measure schemes and its validation in comparison to the ABM model.

In the final step, we demonstrate how to compute Pareto sets of combinations of counter-measures that form optimal compromises between the conflicting objectives that often seem confusing in finding the best strategy, e.g. for decision makers. To this end, we formulate the respective multi-objective optimization problem, and show how to solve it numerically for a realistic test case. We also discuss the meaning of the results, in particular the meaning of the Pareto set for decision makers.

The novelty of this article lies in the following triad:

A. Construct a micro-model that can be used to describe all counter-measures under consideration explicitly and in detail and can be fitted to real-world spreading data,
B. use this micro-model to find a family of macro-models (ideally a single stand-alone ODE model with counter-measure depending parameters) that is able to predict the epidemic spread under all possible combinations of counter-measures, and
C. use this macro-model as the prediction core within the multi-objective optimization problem with conflicting spread-reduction and socio-economic objectives and solve it numerically, i.e., compute the respective Pareto front.

We do not claim that the macro- or micro-model nor the Pareto strategy presented in this article are the best possible or without alternatives. We only claim the novelty of the combined strategy (A)-(C). Recent literature contains a variety of approaches on optimal control of Covid-19 spreading, see, e.g., [19,22,26,29], with the underlying idea being even older and more general [14]. Some parts from these approaches could also be included in the framework presented in this paper, however, we focus on our core ideas for now to make clear where the novelty lies. Further, these other approaches do not contain a micro-model level and describe the consequences of counter-measures rather implicitly by external controls on the ODE level without explicit representation on a micro-level. Moreover, they do not consider the multi-objective perspective. We found a single multi-objective approach [40] (that claims to be the very first multi-objective one), which also does not contain a micro-level and thus represents counter-measures implicitly without being validated against real-world data.

The main goal of this article is to demonstrate that the realization of the combined strategy (A)-(C) and the subsequent derivation of political options for action is practically possible, if micro- and macro-model as well as a mathematical definition of the different objectives can be tailored to the specific situation.

## 2 Micro model: Agent-based model (ABM)

The ABM proposed in [24, 25] models the population of Berlin (approximately 5 million people, 3.6 of which are living inside the city limits) moving about their day, meeting other agents at work, in school, in public transport, at leisure activities or when shopping. The mobility side of the model had been worked on for many years before the outbreak of SARS-CoV-19 and is based on mobile data [33]. The ABM assumes that an infection may occur when an infectious and a susceptible agent are in the same building or vehicle simultaneously.

### 2.1 Basic Structure

As stated above, the ABM takes the contact model from transport modelling. The transport model in question is activity-based, meaning that it contains complete daily activity chains for every agent. These activities have types (for example home, work, leisure, shop), durations and locations. The activity chains are available for a typical weekday, a typical Saturday and a typical Sunday. This means that for every simulated week the weekday model is run five times and then the Saturday and Sunday models are run.

If a contagious agent and a susceptible agent have contact, then the probability of an infection depends on certain factors which are described in Eq. (1). The simulation progresses by discrete timesteps in time *t*. In each step, the probability for agent *n* to become infected at time *t* is described by [34, 35]

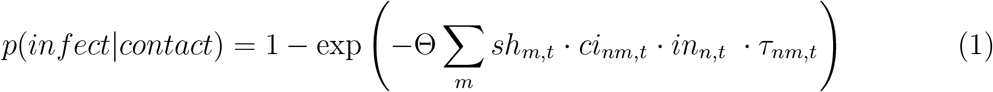

Here *m* is the sum over all agents, *sh* the viral shedding of the infectious agent, *in* the intake, *τ* the duration of the contact, *ci* the contact intensity and Θ an overall scaling parameter. *sh* and *in* can be reduced by wearing a mask, the *ci* depends on the room size and air exchange and *τ* comes directly from the mobility model [25]. All parameters are chosen based on the corresponding literature, and the model is tuned to real-world data, mostly via its single scaling parameter Θ, and to a lesser extent by increasing model detail, for example by including mobility data or outside temperatures, every time when the ever longer case number trajectory can no longer be explained.

#### Aggregation

Every single agent gets a certain infection state, e.g., it is either infected or it is not. Aggregation of all infected agents forms a subgroup. As was hinted at earlier, the infected people are later split into further groups: the exposed, the contagious, the people showing symptoms, the seriously sick and the patients in the ICU. Additionally, one may be interested in the group of the recovered. It is assumed that recovered people are immune. By aggregation, the ABM gives the numbers of people in each group for every day, usually starting in mid-February.

#### Mobility patterns and parametrization

The ABM is strongly data-driven. This section gives a short overview. For details see [25].

- The disease progression model is taken directly from literature and all state transitions follow log normal distributions. The probabilities of requiring hospital or intensive care are age-dependent.
- The disease import from abroad is taken from data publicly provided by the Robert Koch Institute (RKI), the German public agency responsible for collecting this kind of data. This is significant, because the number of imported cases vary strongly depending on the date.
- Since the pandemic started, people decided to reduce some activities or were forced to reduce them due to restrictions. These activity reductions are included in the ABM and are based on mobile phone data.
- In the ABM agents can wear masks depending on the activity types. As the wearing of masks is obligatory in Berlin when shopping and in public transport vehicles this is also included into the model. The compliance rates over time are provided by the local transport company in Berlin (BVG).
- The ABM distinguishes between indoor and outdoor activities. It models the effects of seasonality by assuming that in summer more leisure activities take place outdoors than in winter. If an encounter takes place outdoors, the infection probability is reduced by a factor of 10 compared to the same situation indoors.
- Contact tracing is implemented by putting the contacts of agents that show symptoms into quarantine. The contact tracing is capacity-restricted, meaning that it can only be carried out for a certain number of daily cases. This models that contact tracing can only work effectively when case numbers are low.

### 2.2 Expectation values

Each single ABM simulation is an individual realization of a random process. Therefore, single simulations are of very limited importance. Instead, expectation values over all possible realizations have to be considered. More precisely, assume we consider a vector of *d* observables *x* ∈ ℝ^*d*^ (e.g., for *d* = 5, the aggregated number of exposed, the contagious, the people showing symptoms, the seriously sick and the patients in the ICU), then every random ABM simulation yields *x*(*t*), the respective evolution of the respective numbers in time *t*. The corresponding time-dependent expectation value is denoted

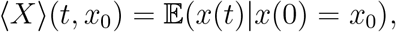

where *x*(0) = *x*_0_ indicates that all realizations start from the same initial state *x*_0_. Its representation as the average over all possible realization implies that ⟨*X*⟩ can be approximated by the running average over a large enough number *m* of independent realizations *x*_*i*_(*t*),

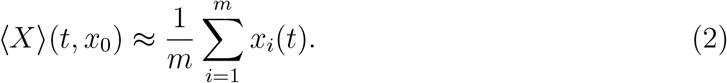

#### Simulation protocol

Each ABM simulation is composed of many individual realizations of the stochastic ABM. Each single one is initialized such that the ensemble of agents can be taken as a realistic synthetic population and such that the respective state of the infection process (including hospitalization etc) is correctly represented.

### 2.3 Counter-measure space

The ABM is flexible enough to model a large variety of counter-measures, ranging from mask wearing (reduced infection probability if more agents wear masks more often) via school closures (changing mobility patterns) to contact tracing procedures (sending agents into self-isolation or quarantine, if exposed). Each single counter-measure *i* can have a weight *c*_*i*_ between *c*_*i*_ = 0 and *c*_*i*_ = 1, representing the strictness of rules and their impact. For example, if *i* represents wearing masks then *c*_*i*_ = 0 means that no-one is wearing them and *c*_*i*_ = 1 means that everyone is wearing a mask all the time.

When measures *i* = 1, …, *M* are considered, every possible combination of counter-measures can be encoded in the counter-measure state *c* ∈ [0, 1]^*M*^ = 𝒞, where 𝒞 is called the counter-measure space. It is important to note that the ABM, if appropriately tuned, allows to perform simulations for all possible states of the counter-measure space. In order to indicate the dependence of ABM simulation results on the respective counter-measure state *c* ∈ 𝒞, we subsequently write respective expectation values by ⟨*X*⟩(*t, x*_0_|*c*).

### 2.4 Validation

The ABM was first fitted to real-world data from Berlin starting February 16, 2020. Figure 1 shows the aggregated simulation results in comparison to real-world observation data published by the Robert Koch Institute (RKI), we see that the aforementioned parameters lead to a simulation result that well reflects the structure of the real data, cf. [30].

**Figure 1:**
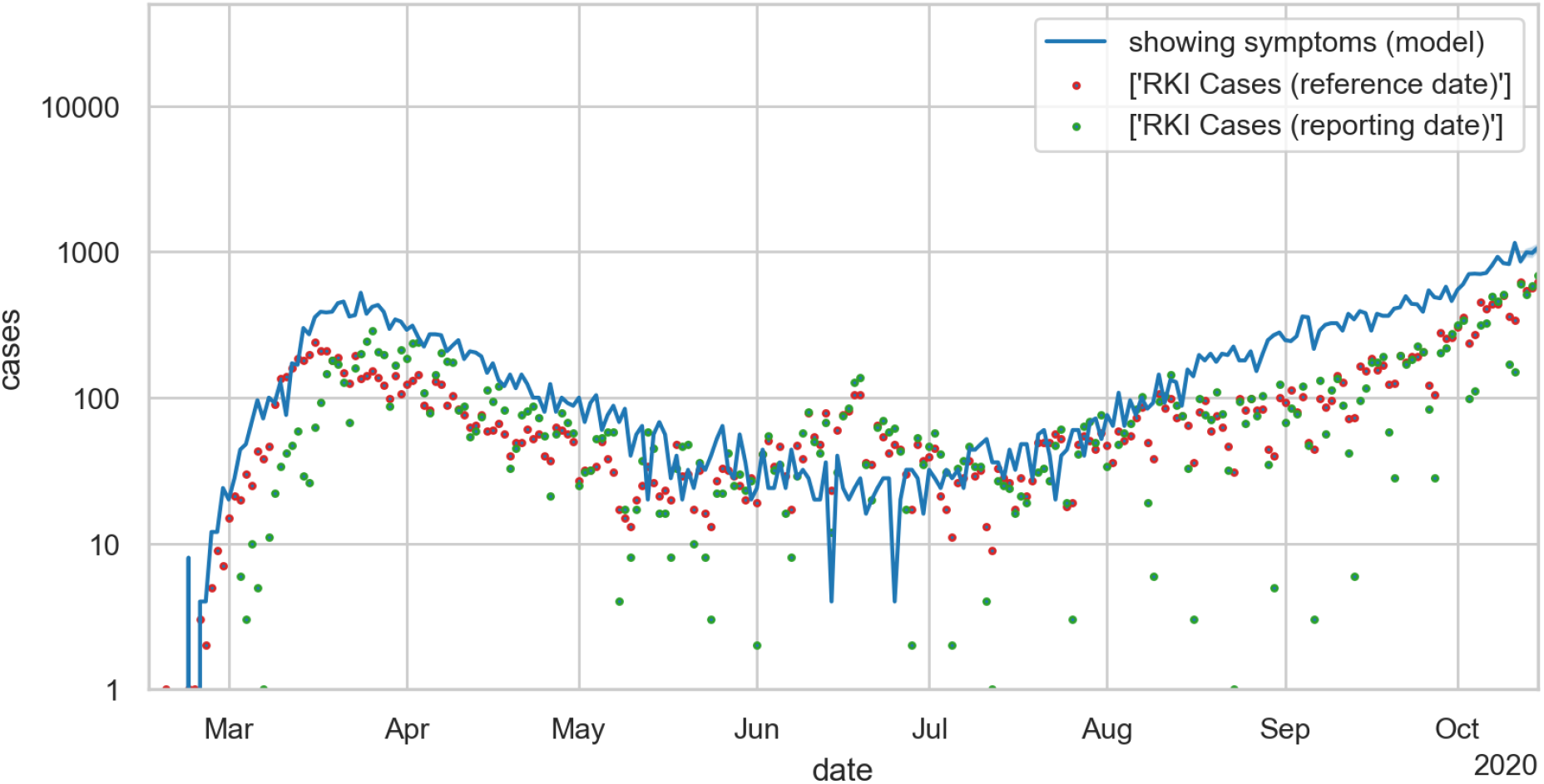
Semi-logarithmic plot of numbers of people showing symptoms for Berlin. Results of a *single* ABM simulation run (blue solid line) in comparison to official RKI observation data (red dots: reference date; green dots: reporting date).

We see that the ABM simulation data reproduces all qualitative trends of the real-world data. In the specific single simulation run shown in Figure 1, the number of people showing symptoms lies above the observed numbers. This is a typical scenario that reflects that not all symptomatic individuals will be recorded by official statistics such that the reported number of symptomatic individuals is slightly under-estimated, cf. [30].

### 2.5 Numerical Experiments: Covid-19 spreading in Berlin

For the sake of simplicity, we only consider three different types of counter-measures (*M* = 3) in this article: school closures, mask compliance and contact tracing. In the following, *c*_1_ = 0 means that schools are open without any restrictions, and *c*_1_ = 1 means that they are completely closed and all classes are online, *c*_2_ = 0 means that no-one is wearing masks and *c*_2_ = 1 means that everyone if earing a mask all the time, *c*_3_ = *γ* means that contract tracing is done for 100 · *γ* percent of the detected infection cases.

#### Simulation data for different counter-measures

The ABM model fitted to real-world data from Berlin for March 01 to October 27, 2020, was used to predict the infection process up to November 30, 2020, for the following combinations of measures for us:

- 0, 50, 100 % school closures
- 0, 50, 100 % mask compliance
- 0, 50, 100 % contact tracing.

This results in 27 different ABM simulation data sets, given as time-series of running averages given by (2) for the aggregation subgroups of interest (given by the compartments required by the ODE model, see below).

## 3 Macro model: Differential equations model (ODE)

A simple model to simulate the spread of infectious diseases such as Covid-19 is the so-called *SIRD* model (***S****usceptible*, ***I****nfectious*, ***R****ecovered*, ***D****eceased*) [13]. This model separates the population of a city/state/country into four *disjoint* groups: Everyone in a population who has not yet been infected by the virus is still *susceptible* to it and is in group S. At the beginning of the pandemic, this was everyone but the first people who got infected (as of yet, it is not clear where the first infections occurred).

From group *S*, people move into the group of the infectious, *I*, with a certain rate. They are also infectious now. The size of this group is important: the more people are already infected, the easier it is for everyone in group *S* to become infected as well. From group *I*, people can either move into group *R* if they have recovered from the virus or into group *D* if they have died.

Usually, the ODEs related to the *SIRD* model are the following:

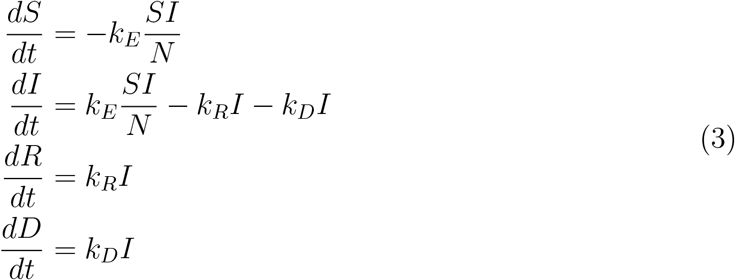

with *k*_*E*_ the infection rate, *k*_*R*_ the recovery rate and *k*_*D*_ the mortality rate.

Because it is so simple, the model has obvious disadvantages, e.g. what about the people who have already been infected but are not infectious themselves yet, or what if the people who are recovered are not actually immune and should be put into the *S* group once more?

There is a variety of ODE models for describing Covid-19 spreading, many of them built on the idea behind the SIRD model but extending it by adding categories, e.g., [19, 22, 26, 29, 40], or by considering other compartmental variants [27]. Other approaches utilize delay differential equations (DDE) [11] or stochastic models, either in the form of stochastic differential equations [41], in discrete form [16] or by probabilistic means [12]. The relation between ABMs and SIRD-like ODE, SDE or DDE models has been discussed from many angles. General results show that the ABM expectation values converge to solutions of respective ODE or SDE models or respective partial differential versions for spatially resolved ABMs when the number of agents is made sufficiently large [17, 37]. However, this condition will not be satisfied in all stages of epidemic spread, especially not in all aggregate classes. Moreover, the theory requires a specific scaling of the stochastic ingredients of the ABM in order to allow for explicit formulas for the ODE and its parameters (which normally will not be available). Therefore, we know that ODE models in general provide appropriate macro-models for ABM micro-models but still need to find ways to fit the macro-model to the specific ABM model at hand.

### 3.1 Description of our ODE model

In order to construct an appropriate ODE model for our purpose, we will extend the basic SIRD model by introducing additional compartments. Our new model will take into account individuals that are exposed, symptomatic, in quarantine and in hospital. A graphical representation of compartments in our model and their relations is shown in Fig. 2. We introduce the following new compartments:

**Figure 2:**
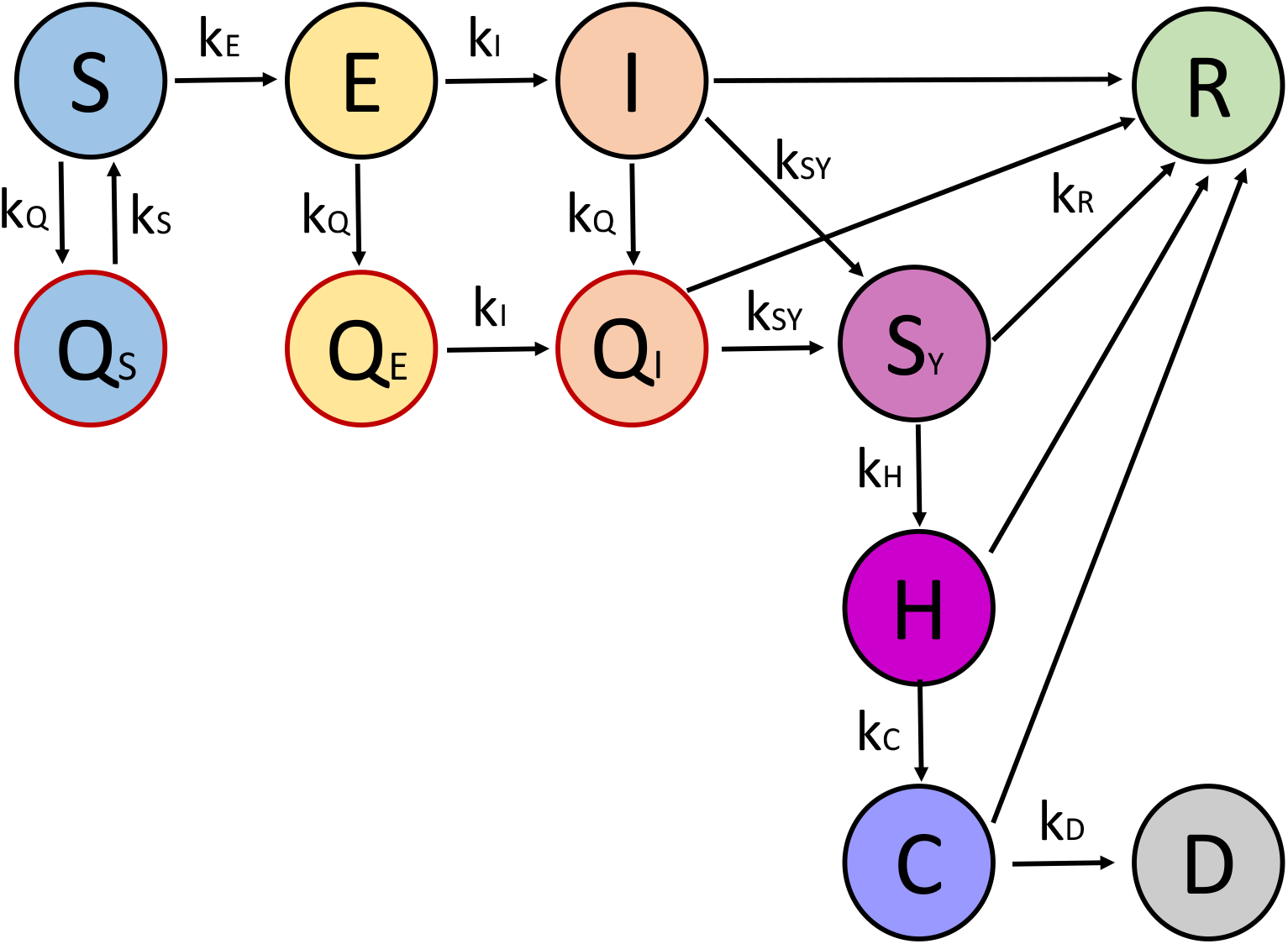
Schematic illustration of our model for the spread of Covid-19 in Berlin.

- **Exposed** *E*: refers to the group of people who have been infected with the virus, but are not infectious themselves yet. After an incubation time, individuals from *E* become infectious and automatically move into group *I*, so in this model it is assumed that individuals do not recover without becoming contagious. A switch from *E* to *I* happens with a rate *k*_*I*_.
- **Symptomatic** *S*_*Y*_: for individuals who are showing symptoms. Infected individuals become symptomatic with a rate *k*_*SY*_. Differentiating between the groups of infectious *I* and symptomatic *S*_*Y*_ individuals is important because it can help in finding undetected cases. In principle, it is more likely that people who are showing symptoms get tested and thus appear in statistics.
- **Hospitalized** *H* **and critical** *C*: refer to patients in hospitals and individuals in critical condition that are in the intensive care unit. We assume that a certain number of people in *S*_*Y*_ recover and directly transit to the recovered compartment *R*. However, the rest is moved into the hospital compartment with a rate *k*_*H*_ and from there, if they recover they go into group *R* or if they are critical they move into the new compartment *C* with a rate *k*_*C*_. From compartment *C*, individuals can go either to *R* or *D*. Please note that the model assumes that people who die from an infection with Covid-19 have been showing symptoms, then were taken to the hospital and into the ICU, i.e. there is now no other way to reach group *D* except from a compartment of critical patients.
- **Quarantine** *Q*_*S*_ **and Self-isolation** *Q*_*E*_, *Q*_*I*_: for individuals that are separated from others by either quarantine or self-isolation. Susceptible individuals who are suspected to have become infected (e.g. by a close contact to a confirmed case) go to quarantine *Q*_*S*_ with a rate *k*_*Q*_. From quarantine, one can go back into the compartment *S* with a rate *k*_*S*_, as *Q*_*S*_ refers to individuals that were not infected and quarantine “in vain”. Exposed individuals *E* and infectious individuals *I* go to self-isolation with a rate *k*_*Q*_, i.e. to compartments *Q*_*E*_ and *Q*_*I*_, respectively. Similarly to the case of *E* → *I*, we also introduce *Q*_*E*_ → *Q*_*I*_ with a rate *k*_*I*_, when self-isolated individuals start to be infectious. Finally, from *Q*_*I*_ possible transitions are either to recovery *R* or to *S*_*Y*_ if symptoms start to appear. These three compartments are necessary to model the effects of contact tracing and related counter-measures.

Thus, the total population is divided into 11 different compartments: susceptible *S*, quarantined *Q*_*S*_, exposed *E*, non-infectious self-isolated *Q*_*E*_, infectious *I*, infectious self-isolated *Q*_*I*_, symptomatic *S*_*Y*_, hospitalized *H*, critical *C*, deceased *D* and recovered *R*. The final model can be described using differential equations, in the following way:

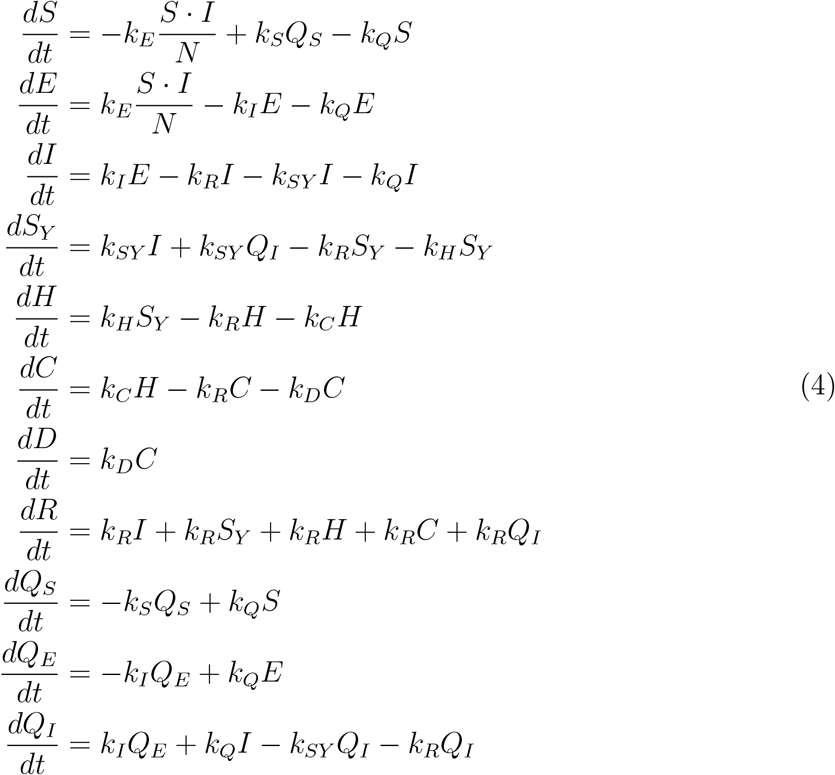

The ODE model contains the following *model parameters*:

- the infection rate *k*_*E*_
- the rate of becoming infectious *k*_*I*_
- the rate of showing symptoms *k*_*SY*_
- the rate of being hospitalized *k*_*H*_
- the rate of becoming a critical patient *k*_*C*_
- the rate of recovering *k*_*R*_
- the rate of mortality *k*_*D*_
- the rate of going into self-isolation/quarantine *k*_*Q*_
- the rate of going out of quarantine *k*_*S*_.

Most of these ODE parameters are dependent on the virus itself and not on any non-pharmaceutical interventions: for example, the rate of being hospitalized should not depend on school closures, but should stay the same no matter the government measures. The only two parameters that should change depending on the measures are the infection rate *k*_*E*_ and the parameter *k*_*Q*_. The latter depends on how well the *contact tracing* is working. Contact tracing usually means the following process: if a person tests positive, they state all the people they have been in contact with over a certain time period. These people are alerted (either by the national health services or by a contact tracing app). If they have been in close contact with the infected person (e.g. by having a long conversation with them or sitting next to them for hours at work), they will be required to quarantine and possibly get tested. If the interaction was not long enough or they kept their distance, they are usually advised to monitor their health and quarantine if they develop any symptoms [3]. Contact tracing is a very effective measure in containing the spread of the virus, but, as has also been worked out in [21, 24], it stops working once the number of cases becomes too high (because health services cannot trace back all contacts anymore). In order to include the effect of contact tracing, we set that

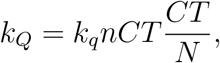

where *k*_*q*_ is the rate of people going into quarantine after being called by the contact tracing agency (*k*_*q*_ = 10.25 independent of counter-measures), *nCT* is the number of people going to self-isolation/quarantine per symptomatic person (a parameter dependent on the effectiveness of the contract tracing agency), and *CT* = *CT* (*t*) is the number of positively tested individuals presently processed by the contract tracing agency. This number is changing with time according to

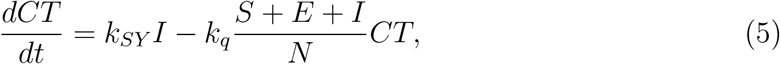

with an inflowing rate given by the number of infected individuals that become symptomatic per unit of time and an outgoing rate with which individuals presently processed by contract tracing go into self-isolation/quarantine and are no longer processed. The number *CT* can approximately be taken as the number of positively tested individuals that are not already quarantined/isolated.

Since *k*_*q*_ is fixed, we have to consider either *k*_*Q*_ or *nCT* as free parameter. Since the definition of *k*_*Q*_ is more involved, we decide to give this role to *nCT*. Additionally, comparable to *k*_*q*_, the parameter *k*_*S*_ turns out to be independent of counter-measures and can be fixed to *k*_*S*_ = 0.192 for all further investigations. Therefore, we will subsequently call the vector *k* = (*k*_*E*_, *k*_*I*_, *k*_*SY*_, *k*_*H*_, *k*_*C*_, *k*_*R*_, *k*_*D*_, *nCT*) the ODE parameters; all possible values of the ODE parameters come from the parameter space 𝒫. The ODE model itself will be denoted

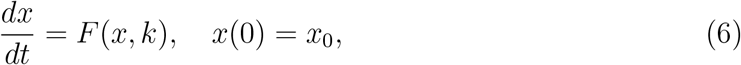

where *x* = (*S, E, I, S*_*Y*_, *H, C, D, R, Q*_*S*_, *Q*_*E*_, *Q*_*I*_, *CT*) ∈ 𝒳 denotes the state vector of all the observables appearing in (4) and (5), and *k* ∈ 𝒫 the respective ODE parameters. The resulting solution of the ODE model is denoted *x*(*t, x*_0_, *k*) for *t* ∈ [0, *T*].

### 3.2 Parameter Estimation

Parameter estimation (PE) means the process of fitting the ODE model to given data. More precisely, one distinguishes between the forward and the backward problem. Generating trajectory data by numerically solving (6) with known ODE parameters is called the *forward problem*. The *backward problem* is the reverse: the data is known but the parameters are not. This is the case here: There is (observation or simulation) data of the spread of the virus; let us denote it by *X*(*t*) ∈ 𝒳, *t* ∈ 𝒯, where 𝒯 is the set of discrete times at which the infection state *x* has been observed. We want to find the ODE parameters for which the ODE solution *x*(*t, x*_0_, *k*) is closest to the given data *X*(*t*) at all time point *t* ∈ 𝒯. In PE for ODEs, closeness is typically measured by means of the relative total *residual* ℛ. The residual depends on the weighted least squares,

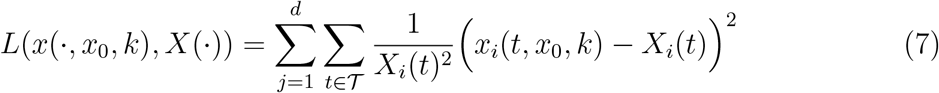

in the following way [18, 31]:

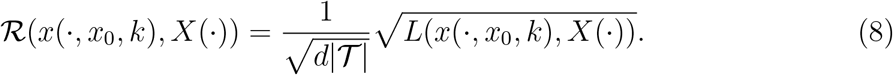

The relative total residual can be defined as the “relative deviation between experiment and simulation per value” [39].

Parameter estimation (PE) means optimally fitting the ODE model to given data *X* by solving the minimization problem [18, 31]

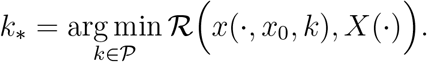

resulting in a non-linear least squares problem. Subsequently, this minimization problem is numerically solved using *PREDICI* [39]. It uses a Gauss-Newton method with damping and reduced directions.

#### Gauss-Newton method and correlated parameters

The Gauss-Newton algorithm is a kind of Newton’s method applied to the first derivative of the vector-valued function *ℒ = ℒ(k*) describing the model-data difference, where its *j*th component, ℒ_*j*_(*k*), is given by the *j* summand in sum in equation (7), *j* = 1, …, *d*|𝒯 |. The key element of the Gauss-Newton method is the rectangular Jacobian matrix *A* of ℒ,

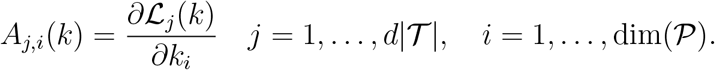

We start with an initial guess for the parameter vector *k*^0^ for the parameters and compute iteratively, using the generalized inverse of the Jacobian,

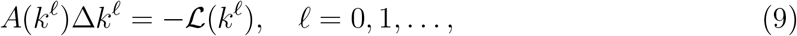

and update the parameter vector due to *k*^.*€*+1^ := *k*^.*€*^ + *λ*_.*€*_ · Δ*k*^.*€*^, where the damping factor *λ*_.*€*_ ≤ 1 is adjusted in each step to ensure improvement of the objective function ℒ.

In many realistic cases, the linear problem (9) is numerically ill-conditioned. The reason for this is that the Jacobian *A* exhibits very small singular values in the vicinity of the solution. These small singular values mean that some of the parameters, say *k*_*i*_ and *k*_*j*_, are *correlated*. This correlation has the effect that ℒ hardly changes if one changes *k*_*i*_ and *k*_*j*_ in a concerted way. Effectively, this means that changes in *k*_*i*_ can be compensated by changes in *k*_*j*_ without ever changing the functional ℒ significantly. The Gauss-Newton algorithm used in *PREDICI* includes a technique that detects parameter correlations during the iteration process and transforms them accordingly. We will see that such correlations are happening when performing parameter estimation for our ODE model which is the main reason why we apply *PREDICI* herein.

It can be shown that the Gauss-Newton method will converge to a local minimum of the residual function. This local minimum is the global one, if a number of conditions are met, one of which is that the initial parameter values must not be too far away from their optimal values: a good first guess of the parameters is crucial to achieving good results in the parameter estimation. Further description of parameter estimation, the underlying minimization problem and its numerical solution as well as the conditions under which it works well, including correlated parameters and choosing initial parameter values, can be found in [18], pp. 230.

### 3.3 Numerical Experiments

Next, we will discuss whether the ODE model is appropriate to allow for reasonably good fit to real-world observation data as well as to ABM simulation data for different counter-measures.

#### Fitting the ODE model to observation data

In order to test how well the ODE model can reproduce real-life data, we will use a set of data for the State of Berlin, including the cumulative numbers of positive tests, the number of hospital beds that are occupied by Covid-19 patients, the number of ICU beds that are occupied by Covid-19 patients and the cumulative number of people who have died after a Covid-19 infection. These data are officially available from March 1 to the moment our experiments were performed, November 5, 2020, through websites fed by the authorities of the State of Berlin. Our model does not have an explicit group that represents positive tests. We identified the group of the symptomatic infected with the group of the positively tested.

Important here: The infection rate kE has to be somewhat time-dependent for real-life data because the infection rate is dependent on the current counter-measures that are in place. In reality, measures and restrictions to prevent the spread of Covid-19 do not come into action all at the same time but are introduced (and lifted) gradually. Sometimes it is only their strength that is varied. If we only have one parameter representing the measures for the complete time span we are looking at, the model will fail to fit real-life data.

So in order for the model to have a chance at fitting the real-life data well, we allow the infection rates to vary with time:

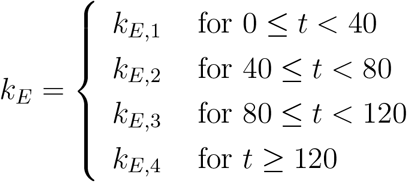

The parameter *k*_*SY*_ is time-dependent in the same way. This is necessary because we equal the number of positive tests to the number of people showing symptoms. The parameter *k*_*SY*_ works as a testing rate and the number of tests have changed significantly over time.

For fitting the ODE model, we use the real-life data from May 18 until October 26, 2020. The data starts at the end of the “first wave” in May, so well before the second wave started taking effect in the fall of 2020. For this period, the parameter estimation scheme introduced above was applied and the resulting optimal parameter values were used subsequently to compute predictions for the infection dynamics beyond October 26, 2020.

The data from October 27 to November 5 (the specific date the numerical experiments were made) were taken as test data for validation of the prediction quality of the model. The result of the optimal fit and resulting prediction/validation is shown in Fig. 3. We observe that the optimally fitted ODE model reproduces the real-world data quite well and yields very good prediction quality.

**Figure 3:**
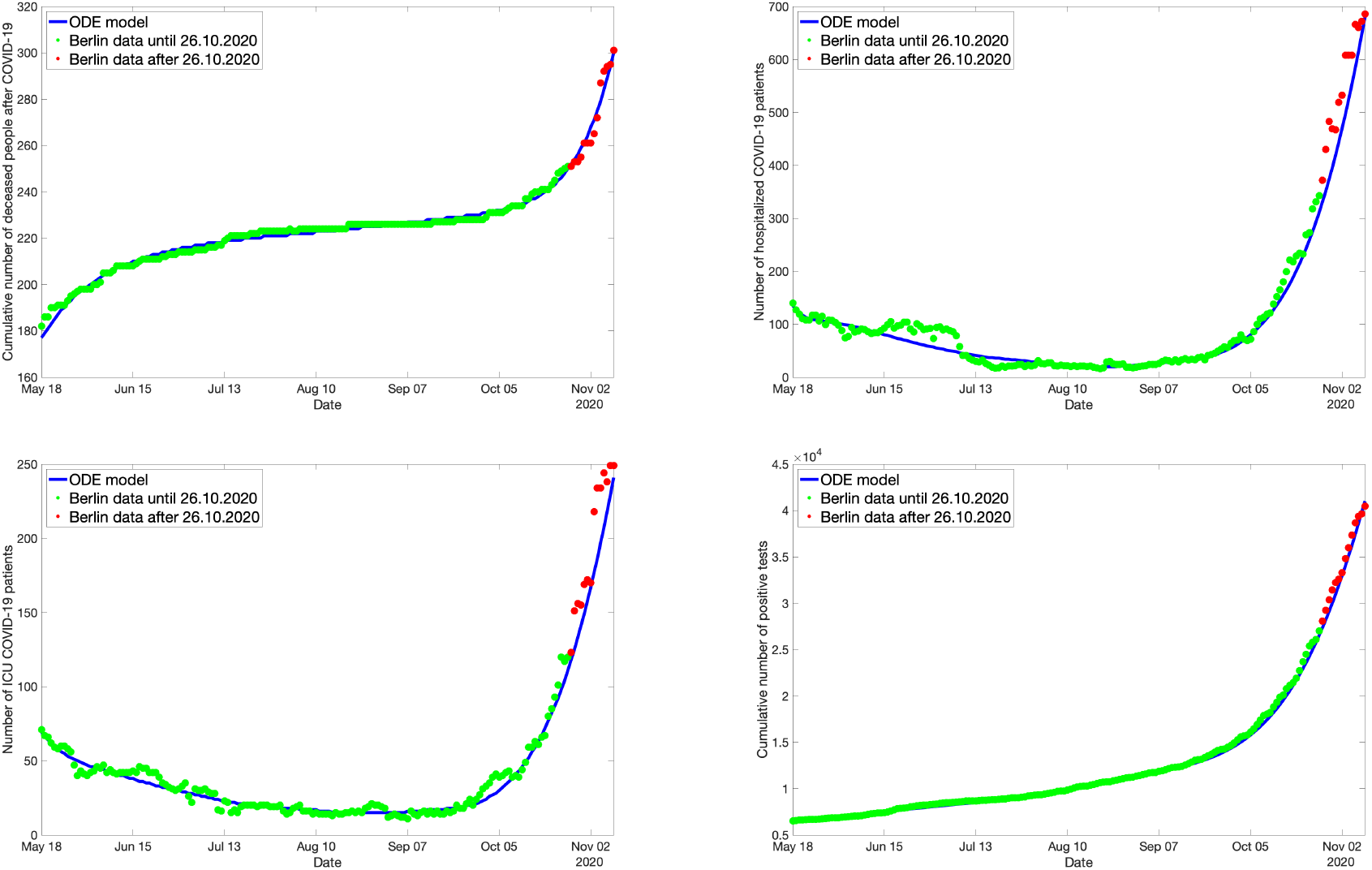
Predictions based on the ODE model for the spread of Covid-19 in Berlin: Cumulative number of deceased (top, left), number of hospital beds occupied (top, right), number of ICU beds occupied (bottom, left), and cumulative number of positively tested persons (bottom, right). Green dots: Real-life data used for fitting the ODE model. Blue solid line: Solution of optimally fitted ODE model. Red dots: Real-life data used for validation.

#### Fitting the ODE model to ABM simulation data

Next, instead of using real-life data to fit the ODE, we use simulation data coming directly from the ABM. This data consists of time-series of aggregated numbers (expectation values) for the model compartments. The initial values are the corresponding real Berlin numbers on the 27th October 2020. The ABM was used to simulate the change in these numbers for the next 5 weeks (until the end of November), assuming certain counter-measures (as outlined in Sec. 2.5).

First, we consider the case where, starting on the 27th of October, all schools (= kindergarten, primary school, secondary school, university, other education) are closed in Berlin, everyone is wearing a surgical mask in public transport, in shops and at work, and contact tracing works faultlessly in identifying each contact of a person that is symptomatic, which is associated with the counter-measure space *c* = (1, 1, 1) according to Sec. 2.5. The ODE fit to this data set is shown in Figure 4. The residual was less than 2 %.

**Figure 4:**
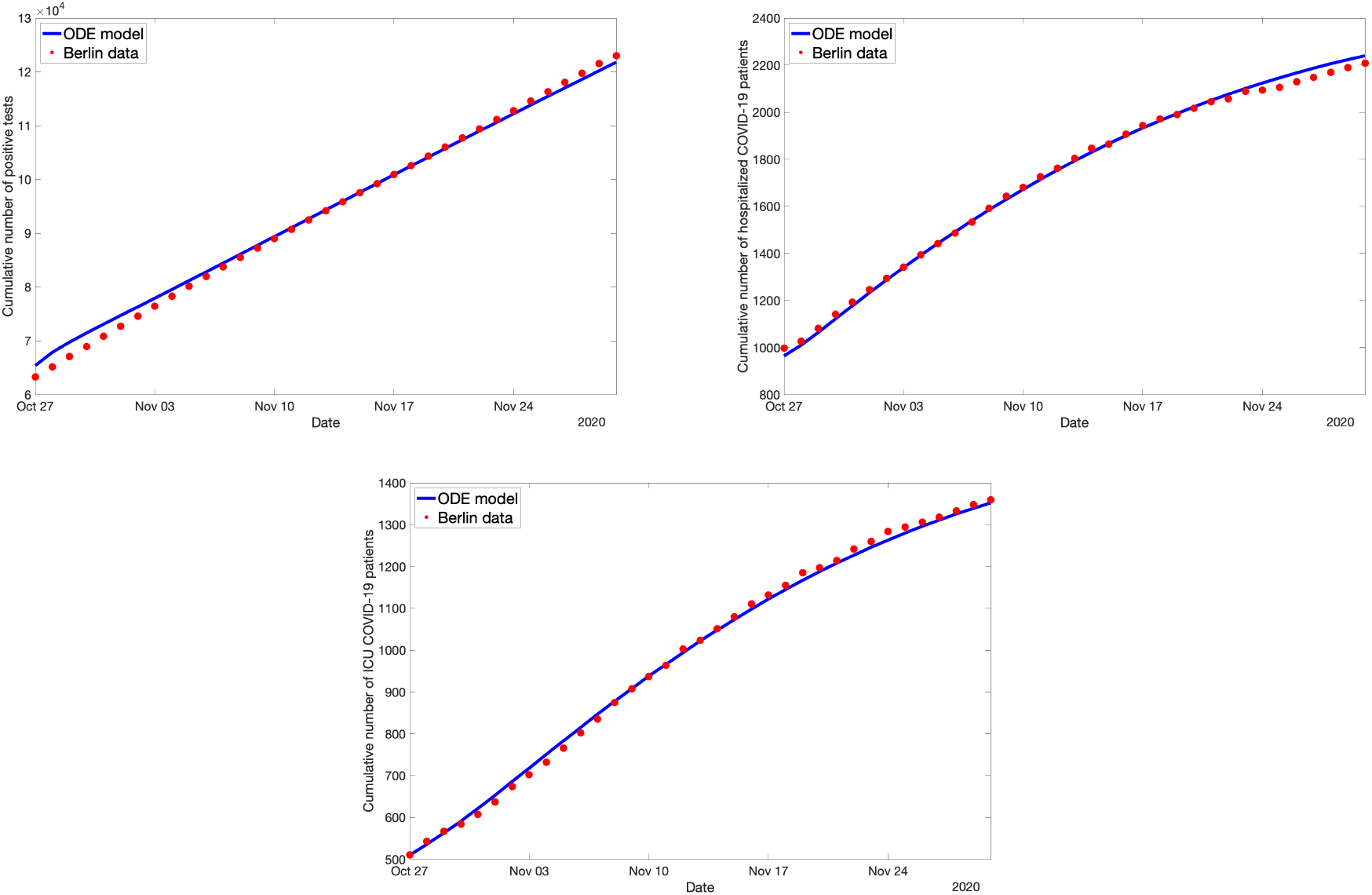
Optimal ODE fit for the ABM data simulating 100 % school closures, 100 % mask compliance and 100 % contact tracing.

Next, consider *c* = (0, 0, 0) starting on October 27 (all schools are open, nobody is wearing masks, no contact tracing is done). Then the ABM simulation data looks quite different, and still the ODE can fit the data nicely, as can be seen in Figure 5.

**Figure 5:**
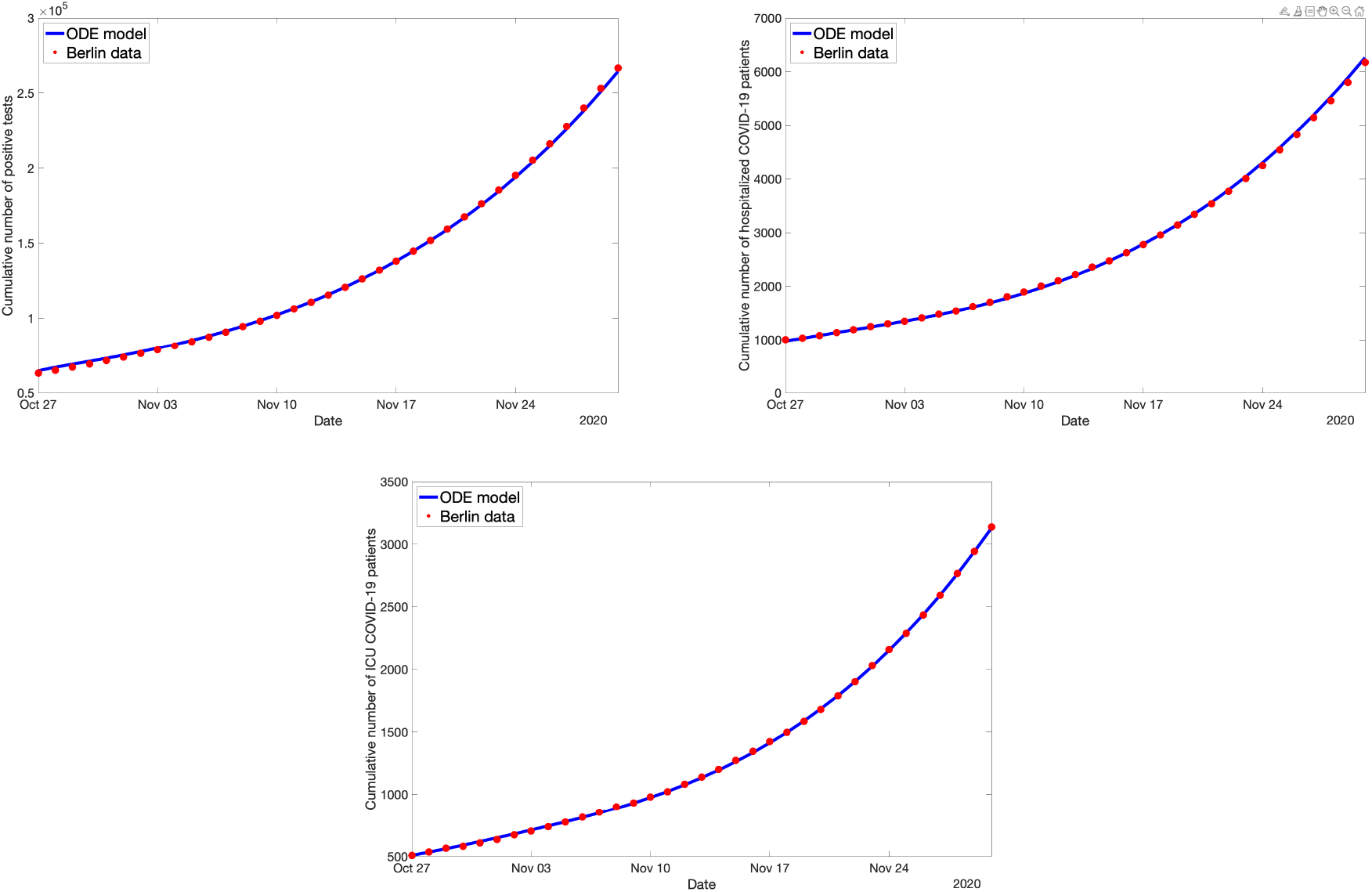
Optimal ODE fit for the ABM data simulating no school closures, no mask compliance and no contact tracing

If we now take a look at the respective optimal ODE parameters as listed in Table 1, we see that except for the infection rate *k*_*E*_ and the contract tracing parameter *nCT* the parameters are roughly the same for both cases.

**Table 1:** Optimal parameter values for the two cases *c* = (1, 1, 1) and *c* = (0, 0, 0) respectively.

| Parameter | $c = (1, 1, 1)$ | $c = (0, 0, 0)$ |
| --- | --- | --- |
| $k_E$ | 0.922 | 1.69 |
| $k_I$ | 0.105 | 0.08 |
| $k_{SY}$ | 0.795 | 0.645 |
| $k_H$ | 0.013 | 0.017 |
| $k_C$ | 0.173 | 0.174 |
| $k_R$ | 0.030 | 0.036 |
| $k_D$ | 0.262 | 0.255 |
| $nCT$ | 11.7 | 9.94 |

This is exactly what we had hoped to see: a change in the counter-measures should not affect the rate of showing symptoms or of being hospitalized but only the rate with which people are infected.

#### Projection onto one parameter

Now, since it seems that the different kinds of counter-measures affect only the two parameters *k*_*E*_ and *nCT* significantly, the idea is to use them to fit all different kinds of counter-measure data sets. The rest of the parameters are fixed to their optimal values for a “base case”, i.e. no counter-measures are in effect. Consider the examples of different counter-measures and the corresponding parameter combinations shown in Table 2. We first observe that the parameters do not behave monotonically, i.e., the infection rate does not automatically drop when the measures become stricter. Also, the parameter values for *nCT* should be close to 0 for the counter-measure combinations without contact tracing but they are not. It seems that *nCT* “absorbs” some of *k*_*E*_’s meaning. This is exactly the scenario of correlated parameters explained in Sec. 3.2: changes in *k*_*E*_ can be compensated by changes in *nCT* without ever changing the residual function significantly. Therefore, in the following we remove this correlation by fitting *k*_*E*_ only, while *nCT* is fixed which effectively means to assume a capacity limit for the contact tracing per day (which makes the model even more realistic). This way, the counter-measure effect of the contact tracing is not visible in the parameter *nCT* anymore but solely in *k*_*E*_, cf. Table 3 below.

**Table 2:** Optimal parameters *k*_*E*_ and *nCT* for different counter-measure combinations while fitting only these two parameters. Obviously, *k*_*E*_ and *nCT* are highly correlated.

| Counter-measure combination | $k_E$ | $nCT$ |
| --- | --- | --- |
| $c = (0, 0, 0)$ | 1.658 | 13 |
| $c = (1, 0, 0)$ | 1.52 | 7 |
| $c = (1, 0.5, 0)$ | 1.64 | 23 |
| $c = (0.5, 0.5, 0.5)$ | 1.849 | 78 |
| $c = (0.5, 0.5, 1)$ | 2.035 | 124 |
| $c = (1, 1, 1)$ | 2.19 | 169 |

**Table 3:** Parameter *k*_*E*_ for different counter-measure combinations. The respective optimal fit to the different data sets exhibits low residuals in all cases.

| Counter-measure combination | $k_E$ |
| --- | --- |
| $c = (0, 0, 0)$ | 1.24 |
| $c = (1, 0, 0)$ | 1.14 |
| $c = (1, 0.5, 0)$ | 1.1 |
| $c = (0.5, 0.5, 0.5)$ | 0.965 |
| $c = (0.5, 0.5, 1)$ | 0.8886 |
| $c = (1, 1, 1)$ | 0.8287 |

With this idea, we fit the base case *c* = (0, 0, 0) using all parameters again, fix all of them except for *k*_*E*_ and compute the optimal fit to the case *c* = (1, 1, 1) using only *k*_*E*_. The result is shown in Fig. 6. While the fit is not as good as it was when all parameters were fitted, the residual here is less than 3 %, so still very small.

**Figure 6:**
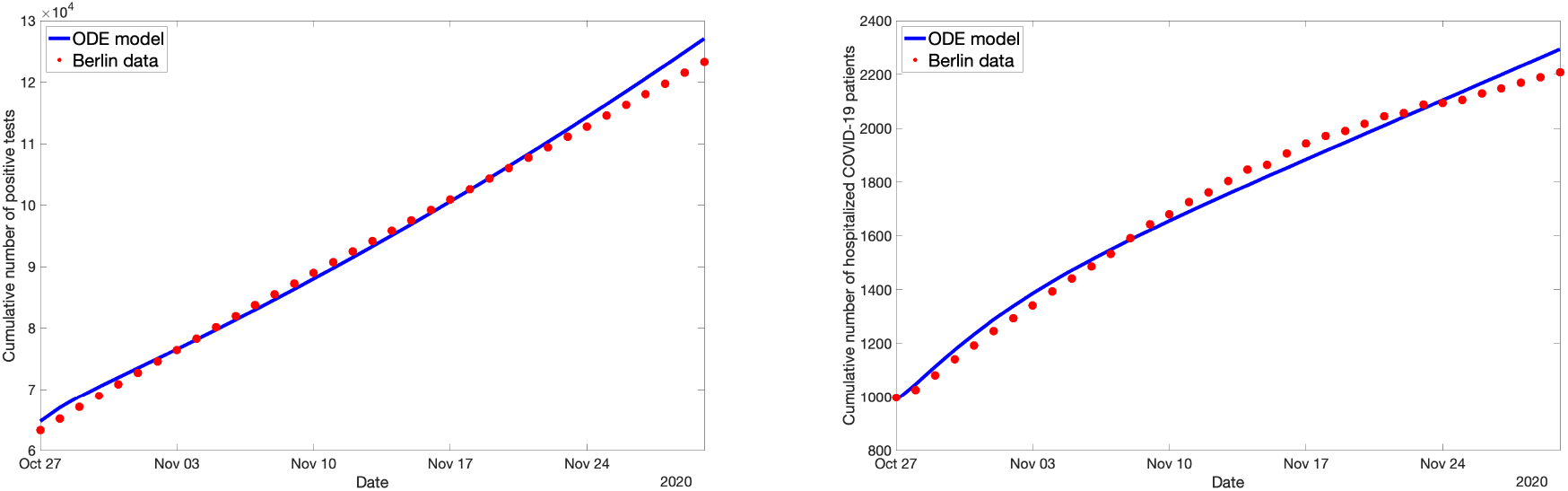
The ODE fit for the ABM data simulating 100 % school closures, 100 % mask compliance and 100 % contact tracing. This time, only *k*_*E*_ was used to fit the data.

Now, when changing the counter-measure state *c* the optimal parameter value of *k*_*E*_ does decrease monotonically with stricter measures, as can be seen in Table 3.

### 3.4 Stand-alone ODE model

In principle, the ABM simulation can be performed for all states in the counter-measure space 𝒞 (keeping in mind that each simulation causes considerable computational effort). We perform such simulations for *N* counter-measure states *c*_*i*_ ∈ 𝒞, *i* = 1, …, *N*, e.g. from a grid discretization of 𝒞. Then we use PE to fit the ODE model to this simulation data, i.e., find optimal ODE parameters *k*_*∗*_(*c*_*i*_), and learn a function *K*: 𝒞 → 𝒫 from this parameter data. For unknown counter-measure states *c*′, we then use the ODE model with ODE parameters *K*(*c*′). When successful, we can now use the ODE model as a stand-alone model

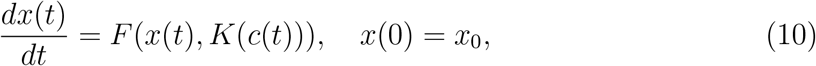

for every counter-measure state *c* that may even depend on time *t*.

More precisely, the proposed procedure for learning the stand-alone ODE model works as follows:

Parametrization and validation of the stand-alone ODE model:

A. Generate ABM training data: For each counter-measure state *c*_*i*_, *i* = 1, …, *N*, perform *m* independent ABM simulations all starting from the same initial state *x*_0_; denote the resulting expectation value wrt. the chosen observables *x* ∈ 𝒳 by ⟨*X*⟩(*t*|*c*_*i*_).
B. Fitting the ODE model to the ABM training data: For each counter-measure combination *c*_*i*_, fit the ODE model to the simulation data ⟨*X*⟩(*t*|*c*_*i*_) using the PE algorithm and identify optimal parameters *k*_*∗*_(*c*_*i*_) ∈ 𝒫. Check whether the residual *r*_*i*_ = ℛ(*x*(·, *k*_*∗*_(*c*_*i*_)), ⟨*X*⟩(·|*c*_*i*_)) is comparably small for all *i* = 1, …, *N*.
C. Use nonlinear regression to find an optimal fit function *K*: 𝒞 → 𝒫 on counter-measure space 𝒞 to the data *k*_*∗*_(*c*_*i*_), *i* = 1, …, *N*.
D. Generate ABM testing data: For each counter-measure combination *c*_*i*_, *i* = *N* + 1, …, *N* + *N* ′, perform *m* independent ABM simulations; denote the expectation values of these simulations on the observable space 𝒳 by ⟨*X*⟩(*t*|*c*_*i*_).
E. Validation: For each of the *N* ′ counter-measure combination *c*_*i*_, *i* = *N* +1, …, *N* + *N* ′, compute the residual

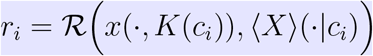

of the ODE model wrt. the ABM testing data.

Remark: The number *N* + *N* ′ of required independent sets of ABM simulation data must not be too large because of limitations of available computational capacities. In particular, this means that for dimensions of the counter-measure space larger than 3 or 4 one needs to resort to sparse grids or quasi Monte-Carlo methods for choosing the counter-measures states in Step A.

In the context of the infection spreading in Berlin described above, these five steps can be realized as follows.

#### Step A: ABM testing data

We put this procedure to practice following the scheme described in Sec 2.5. The ABM is first tuned to real-world data from Berlin. Then, starting on October 27, three of the counter-measures are varied in strength: the percentage of schools that are closed; percentage of people who are wearing surgical masks; and the probability that contact tracing is successful. These three counter-measures are adjusted to 0, 50 and 100 % respectively, resulting in 27 different measure combinations that define a uniform grid in counter-measure space, 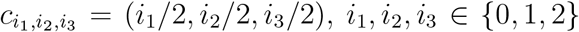. These grid points are used for step (1) in our procedure for defining the stand-alone ODE.

Please note: Because 100 % successful contact tracing is highly unlikely (practically impossible), we assume that 60 % successful contact tracing is the best we can hope for and is represented by *c*_3_ = 1.

Between the most strict set of measures (100 % of schools are closed, everyone is wearing a mask, contact tracing works faultlessly) and the most loose measure set (0 % for all three measures), the simulations show a large difference in the numbers in the projections for the hospital numbers for November (see Figure 7).

**Figure 7:**
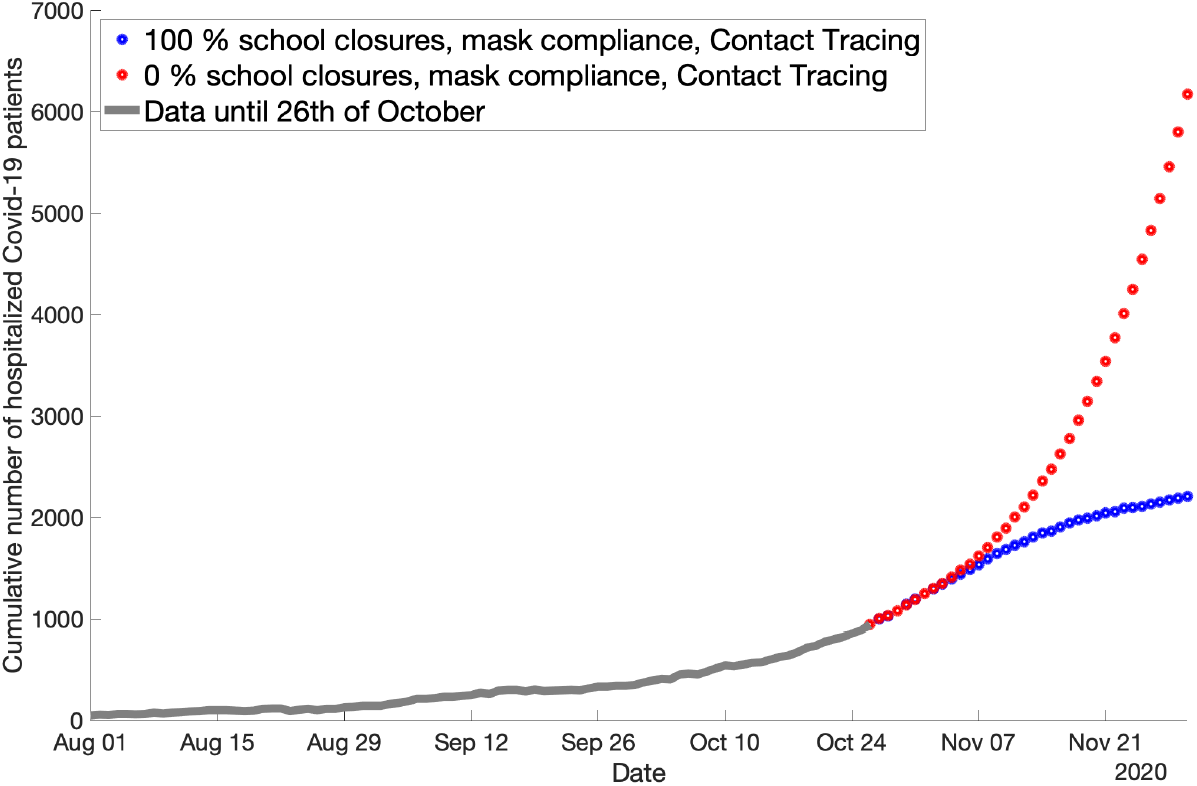
Difference in the ABM predictions of hospitalization numbers in Berlin when different counter-measures are used. The grey line shows the underlying observation data. On October 27, the difference in the counter-measures takes effect. The blue line (*c* = (0, 0, 0)) and the red line (*c* = (1, 1, 1)) show the resulting different projections for November.

#### Step B: Fitting the ODE model

Next, following step (2) in our procedure, we fit the ODE to all of these 27 data sets. Again, we only need to use the infection rate *k*_*E*_ to fit all 27 data sets while fixing the rest of the parameters to a value coming from the optimal fit of a so-called “base case”, in our case the data set with the lowest measures (*C* = (0, 0, 0)). For all 27 data sets, the resulting residuals are very small (less than 5 %). The parameter values that were used can be found in Table 4, except for *kE* and *nCT*. The latter one was fixed to the value of 60, which gave the best results.

**Table 4:** Optimal parameter values for the base case *c* = (0, 0, 0) that are used to determine *k*_*E*_ for the rest of the cases.

| Parameter | Value |
| --- | --- |
| $k_I$ | 0.095 |
| $k_{SY}$ | 0.61 |
| $k_H$ | 0.016 |
| $k_C$ | 0.18 |
| $k_R$ | 0.045 |
| $k_D$ | 0.26 |

#### Step C: Regression on counter-measure space

Until now, we got the values 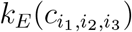 for all grid points 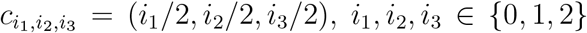 in counter-measure space. Next, we will predict the value of *k*_*E*_ for unknown *c*, without using new and computationally very expensive ABM simulations. To do this, we use polynomial regression to express the function as a polynomial of low degree by computing its coefficients. The polynomial regression is done using the *sklearn* package in Python. As an example, a three-dimensional (i.e. three input values) polynomial of degree 3 has the following form:

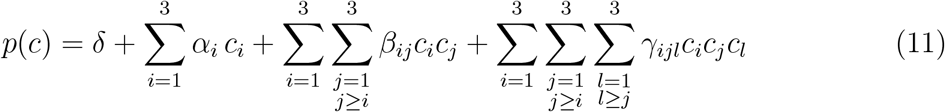

with *δ* denoting the intercept, and *α*_*i*_, *β*_*ij*_, *γ*_*ijl*_ the coefficients to be computed. Hence, this regression model has 20 parameters (19 coefficients plus the intercept). For a polynomial of degree 2 the three sums on the right hand side are omitted.

In the case considered herein, polynomial regression works exceptionally well for a polynomial of degree 2 for *k*_*E*_. We tried three different polynomials, one with degree 2, one with degree 3 and one with degree 4. The respective relative error (compared with the training label sets) are 0.00058, 0.00028 and 0.00017. As can be seen in Figure 8, even though the fit *is* better for a higher-degree polynomial, the error reduction with higher degree does not weigh up the disadvantages of having many more parameters in the regression function which often leads to overfitting on the training set. Therefore, we choose to keep the function with a 2-degree polynomial.

**Figure 8:**
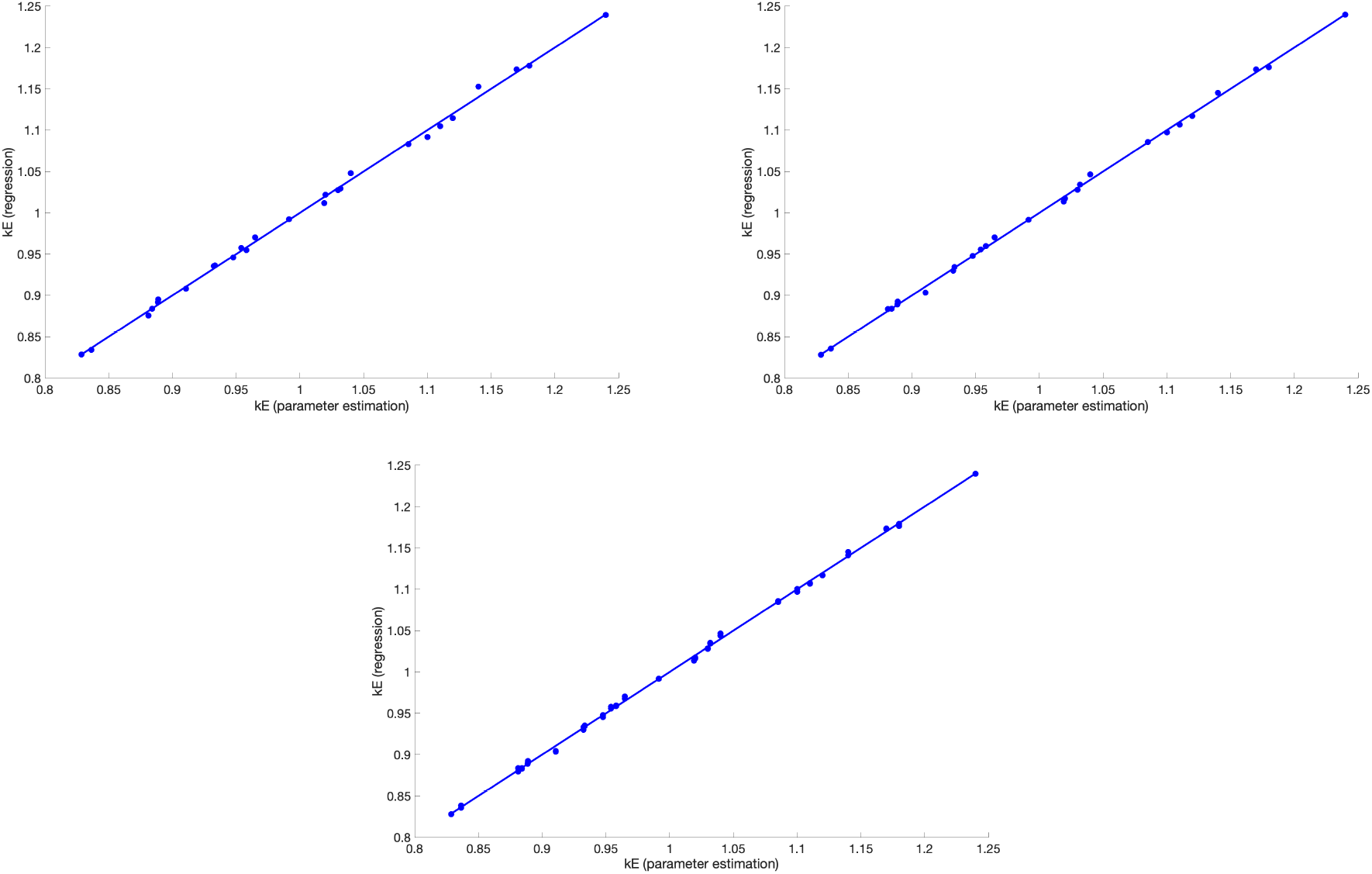
Polynomial regression function for *k*_*E*_ with different degrees. The plots show the result when polynomials of degree 2, 3, or 4, respectively, are used. The accuracy does not improve significantly if a higher degree than 2 is used.

#### Steps D and E: Test data and validation

Next, we first generate ABM testing data for the counter-measure states *c* = (*z/*100, 0.5, 0.5) (“Schools”), *c* = (0.5, *z/*100, 0.5) (“Masks”) and *c* = (0.5, 0.5, *z/*100) (“CT”) with *z* = 10, 20, 30, 40, 60, 70, 80, 90. We compute the value of *k*_*E*_ using the regression function. Afterwards, we insert the resulting values into the ODE model and compute the residual between the ODE model output and the ABM data set. The resulting residuals can be found in Table 5. The largest residual for the validation set is 0.065. This is really acceptable.

**Table 5:** Residuals for the regression parameter predictions for the ABM testing data. The head of each column gives the counter-measure which is varied (see text), i.e. the other two measures are fixed to 50 %.

| z | Schools | Masks | CT |
| --- | --- | --- | --- |
| 10 | 0.026 | 0.046 | 0.065 |
| 20 | 0.031 | 0.038 | 0.048 |
| 30 | 0.025 | 0.031 | 0.041 |
| 40 | 0.027 | 0.03 | 0.03 |
| 60 | 0.029 | 0.027 | 0.03 |
| 70 | 0.028 | 0.027 | 0.03 |
| 80 | 0.034 | 0.029 | 0.032 |
| 90 | 0.031 | 0.025 | 0.027 |

### 3.5 Predictions using the stand-alone ODE model

Now that the regression results have been validated, we can use them to make predictions on the effectiveness of counter-measure combinations. Consider the following examples.

#### Example 1

Using the regression results, we predict that the infection rate for the counter-measure combination *c* = (0.99, 0.50, 0.59) should be *k*_*E*_ = 0.936. In this scenario, schools are almost completely closed.

Now, for the counter-measure combination *c* = (0.16, 0.50, 0.73), the infection rate *k*_*E*_ = 0.936 as well. This means, given that we trust the regression results to a certain extent, the effect of these two counter-measure combinations are the same. It also shows that the effect of school closures are rather small in the ABM, which fits the current research that says that schools are not driver of the pandemic, at least in Germany.

#### Example 2

As another example, the regression approximation of the infection rate for *c* = (0.12, 0.15, 0.7) yields *k*_*E*_ = 1. This is also the resulting infection rate for *c* = (0.9, 0.9, 0.2). Again, we see that contact tracing compensates for a drastic reduction in the number of schools closed and masks worn.

Now that we can approximate which counter-measure combinations lead to which infection rate, we can tailor the optimal restriction response in order to achieve a certain goal, for example to never exceed a certain number of hospitalized Covid-19 patients as to not overwhelm hospitals in Berlin. There are certain measures that are seen to be less intrusive in people’s daily lives and should thus be used preferentially, as long as they lower the infection numbers sufficiently. In the first example above, we see that we only need to use a bit more contact tracing to compensate for opening the schools almost completely (16 % closed = 84 % open). This would then be seen as the preferred course of action, as contact tracing does not affect society as much as closing schools (which in turn leads to parents not being able to work etc.).

## 4 Optimal coordination of counter-measures

For an optimal coordination of counter-measures one first has to define the objectives with respect to which optimality has to be judged. For the Covid-19 pandemic there will be many such conflicting objectives: on the one hand, decreasing the number of infected, or, at least, keeping the number of deaths minimal and the number of ICU beds below a certain threshold given by the capacity of the health care system; on the other hand minimizing socio-economic, political and/or health care system costs. Let us assume that the relevant objectives can be modelled by a vector of *n* objective functions

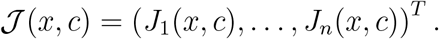

Additionally, we have to define the set of all feasible counter-measure schemes. A counter-measure scheme is a function *C*: [*t*_1_, *t*_2_] → 𝒞 that defines a counter-measure state for each time *t* in the time interval [*t*_1_, *t*_2_] in which we are looking for an optimal combination of counter-measures. The feasible counter-measure schemes have to satisfy certain restrictions: one cannot switch too often between counter-measure states and between switches the counter-measure state has to be constant. Let the set of feasible counter-measures be denoted by 𝒞[*t*_0_, *t*_1_].

The objective functions *J*_*k*_(*x, C*) depend on the counter-measure scheme *C*: [*t*_1_, *t*_2_] → 𝒞 and the associated infection dynamics *x*: [*t*_1_, *t*_2_] → 𝒳, in general in the form

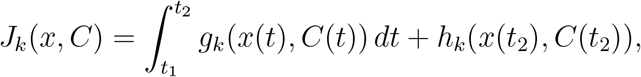

where the terminal cost function *h*_*k*_ or the running cost *g*_*k*_ may be zero. Then, an optimal counter-measures scheme *C*_*∗*_(*t*) ∈ 𝒞[*t*_0_, *t*_1_] is given by a solution of the multi-objective optimization problem

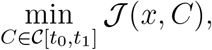

subject to the infection spread dynamics given by the stand-alone ODE model

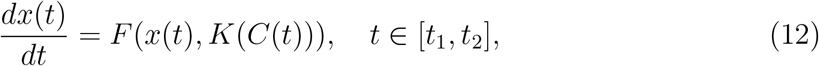

fitted to ABM simulation data in the time period [0, *t*_1_].

For the sake of simplicity of notation, we now write our optimization problem in the following form:

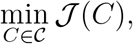

suppressing for the moment that *C* is a time-dependent function, or assuming that the space 𝒞[*t*_0_, *t*_1_] is a finite-dimensional function space.

In such a multi-objective optimization problem, there does not typically exist a feasible counter-measure scheme that minimizes all objective functions simultaneously. Therefore, one seeks to find Pareto optimal counter-measure schemes; that is, schemes that cannot be improved in any of the objectives without degrading at least one of the other objectives. Subsequently, we will write 𝒥 (*C*) ≤_*p*_ 𝒥 (*C*′) if *J*_*k*_(*C*) ≤ *J*_*k*_(*C*′) for all *k* = 1, …, *n*. A scheme *C* ∈ 𝒞 is called Pareto-dominant if there is no other scheme 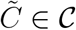 such that

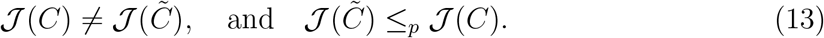

A counter-measure scheme *C* ∈ 𝒞 is called (globally) *Pareto-optimal* if there does not exist another scheme that Pareto-dominates it. The set of Pareto optimal schemes is often called the *Pareto front*. According to [20], each Pareto-optimal scheme is a sub-stationary scheme. A scheme *C* ∈ 𝒞 is called sub-stationary if there is a sequence of non-negative scalar weights 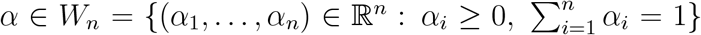, such that

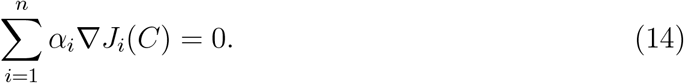

The condition in (14) means that the weighted objective (“compromise objective”)

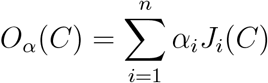

has a local minimum at the sub-stationary scheme *C*. The set of sub-stationary schemes *C* ∈ 𝒞 typically forms a (*n*-1)-dimensional manifold in 𝒞 which contains the Pareto front; in this sense, the set of sub-stationary schemes represents all possible optimal compromises (resulting from minimizing all *O*_*α*_ for *α* ∈ *W*_*n*_).

### 4.1 Numerical Experiments

In order to keep the numerical experiments simple, we consider only two objectives: the number of symptomatic infected, and the socio-economic costs, quite over-simplifiedly modelled by a linear function of *c*,

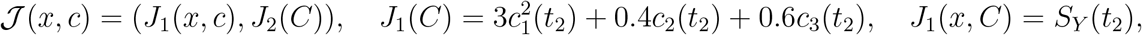

where we chose to consider terminal costs at the end time *t*_2_ only. The choice of *J*_1_ means that the costs related to high percentages of school closures are estimated about several times higher than that of high percentages of mask wearing and contract tracing. Moreover, the costs related to contract tracing are estimated about 50% higher than those of mask wearing.

Moreover, we define the set of feasible counter-measures by

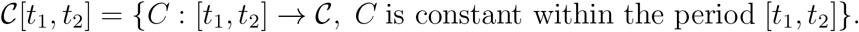

Thus, 𝒞[*t*_1_, *t*_2_] can be identified with [0, 1]^3^.

#### Numerical approximation of the Pareto front

Based on the theoretical insights on Pareto sets and sub-stationary points outlined above, the authors of [10] propose a multilevel subdivision technique that allows to find a numerical approximation of the Pareto front, also cf. [32]. The main idea is that the solutions of the ordinary differential equation

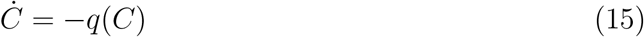

asymptotically converge to sub-stationary schemes, where the function *q* is defined by

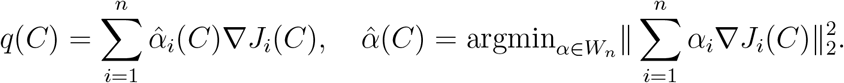

We utilized this algorithmic scheme by starting from a coarse uniform grid *G* on [0, 1]^3^, selected all grid points for which condition (13) is satisfied within *G*, and used this as a start box covering (“level 1”) of the Pareto set, which then was used to start the subdivision algorithm based on the contraction mapping given by (15).

Figure 9 shows the resulting box coverings on level 1 and 3 of the subdivision process. We observe that the Pareto front seems to form a straight line from about (0.1, 1, 0.6) to (0.1, 0, 0.8), approximately. Fig. 10 shows the values of the two objectives plotted against each other along this level 3 covering.

**Figure 9:**
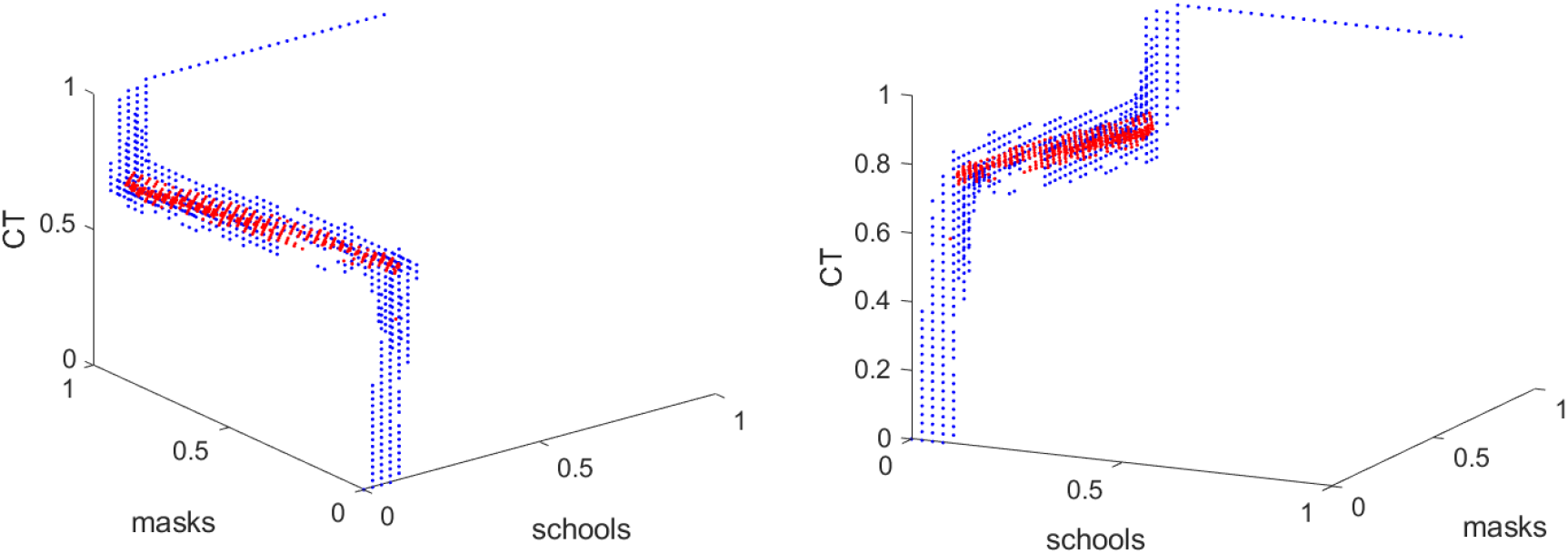
Box covering of Pareto front shown from two different perspectives. Only the box centers are displayed (level 1 (blue dots) and level 3 (red dots)).

**Figure 10:**
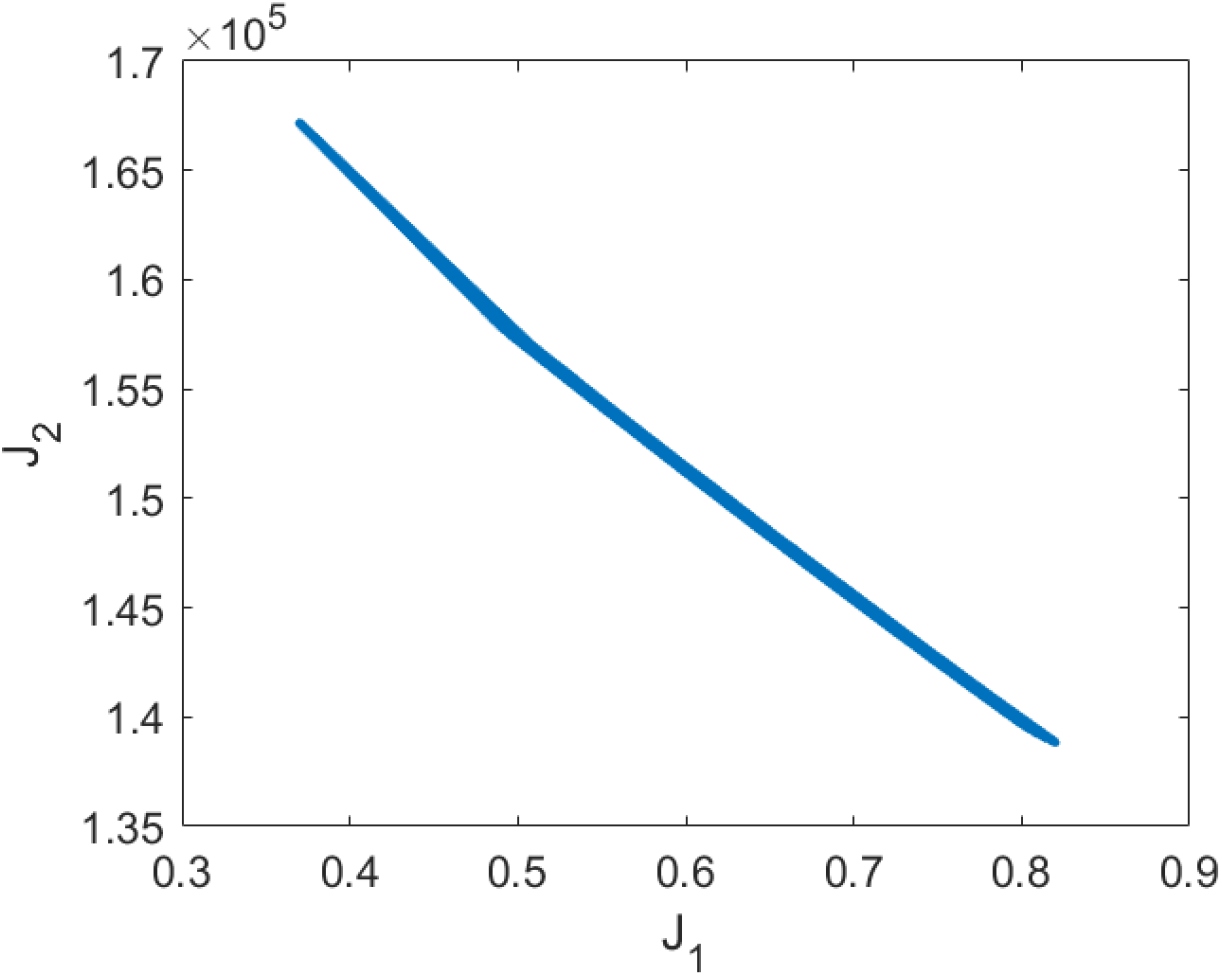
Values of the two objective functions *J*_1_ and *J*_2_ along the Pareto front displayed in Fig. 9 plotted against each other.

We see that *c*_1_ = 0.1 along the Pareto front, approximately, showing that the effect of school closures on suppressing growing infection spread is limited according to the model. Essentially, the Pareto front is spread from *c*_2_ = 0 to *c*_2_ = 1, while *c*_1_ only varies from 0.6 to 0.8. This shows that the percentage of wearing masks is the main variable if one wants to choose one point on the Pareto front, while, whatever is chosen wrt. to masks, contract tracing always has to be quite extensive.

#### Approximately given objective functions

In our case, we use the ODE model as a surrogate for the (more accurate) ABM model. That is, the objective functions *J*_.*€*_ = *J*_.*€*_(*x, C*) are not given precisely but, through the approximate nature of *x* as the solution of the stand-alone ODE, only in a somewhat perturbed form 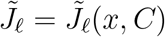. Therefore, we should ask in which sense the Pareto front computed for 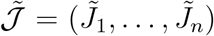 approximates the Pareto front of 𝒥 = (*J*_1_, …, *J*_*n*_). This important question is addressed in [28] for the case that a complex micro-model is replaced by a surrogate model. The authors of [28] show how to adapt the subdivision algorithm used above can to this situation by introducing the concept of *ε*-perturbations into the definition of Pareto dominant points for 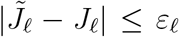 We will not discuss this aspect herein further, mainly because it is unclear how to estimate the *ε*_.*€*_ when replacing the ABM by the ODE model. This topic requires significant further research.

### 4.2 Pareto front based policy decisions

Mathematically, we cannot decide which of the points on the Pareto front is superior or preferable; they all share the unimprovability property of Pareto points: you cannot improve a single objective without making the other ones worse. Any decision about the selection of one of the Pareto points must be based on additional aspects and thus is “political” in the following sense: the fact that there is a Pareto front with more than just one point leaves *freedom for policy decisions*. By choosing one of the Pareto points, the political decision makers execute this freedom wrt. choosing one of many optimal compromises between the conflicting objectives.

In the example above, for example, the specific form of the Pareto front tells us that one is free to decide about the percentage of mask compliance, while there is no freedom regarding the level of school closures, where the percentage of about 10% is given by the Pareto front as optimal. The choice regarding the strength of contact tracing is also limited to between 60 and 80 percent (but only in accordance with the choice for mask compliance.

## 5 Discussion

Micro-level models of epidemic spreading processes allow for the realistic combination of models for person-centric data-driven human mobility and behavior, stochastic infection models and person-centric disease progression models with governmental intervention strategies. This allows detailed simulations of the societal transmission from government actions to mobility behavior to infection dynamics. Therefore, they *in principle* would be the perfect basis for predicting the outcome of all possible intervention strategies and then optimize certain objectives over them.

Such micro-level models, however, are much too complex (computationally demanding) to allow for direct integration to multi-level optimization algorithms. Therefore, we demonstrated how to construct a macro-model as a surrogate for the macro-model, in the sense that the macro-model can be fitted to approximately reproduce the dynamical behavior of the micro-model for all intervention strategies (=combination of counter-measures) of interest. Then, using the macro-model in the inner core of a multi-objective optimization solver allows to compute the Pareto front of optimal compromises between the objectives of interest.

By this strategy, considering the control of the spreading of the diseases as an optimization problem can help governments choose strategies that prevent the disease outbreak as well as limit economic damage.

The results presented in this article demonstrate that this strategy can be realized theoretically and practically but they do not solve all immanent problems. Several important aspects need significant refinement and extended future research. For example, (1) the mathematical formalization of the objective functions modeling the socio-economic and health system costs is naive and has to be refined based on extended non-mathematical expertise, (2) if specific considerations of age or other social aspects in the population or specific test strategies are required, the ODE model must (and can) be extended, (3) the approach can be extended into a network model by taking the exchange with other regions with a different infection scenario into account, (4) the algorithm used for solving the multi-objective optimization problem must be developed further in order to be able to handle more complicated problems.

In conclusion, the present article lays the groundwork for simulation-based real-world multi-objective policy design.

## Data Availability

All data is available through public sources, see refs [1] to [7], and [30].

https://covid-sim.info/

## Acknowledgements

We thank Michael Wulkow for his collaboration on the ODE model and its parametrization using *PREDICI*, and Michael Dellnitz for joint discussions regarding the Pareto front and its numerical approximation.

The work on this paper was funded by the German Ministry of research and education (BMBF) (project ID: 01KX2022A) and by the Deutsche Forschungsgemeinschaft (DFG, German Research Foundation) under Germany’s Excellence Strategy via MATH+: The Berlin Mathematics Research Center (EXC-2046/1, project ID: 390685689).

## References

[1] Blavatnik School of Government. https://www.bsg.ox.ac.uk/research/research-projects/coronavirus-government-response-tracker, accessed September 2020.

[2] Johns Hopkins University. https://coronavirus.jhu.edu/map.html, accessed September 2020.

[3] Robert Koch Institut. https://www.rki.de/DE/Content/InfAZ/N/Neuartiges_Coronavirus/Kontaktperson/Management.html, accessed September 2020.

[4] The COVID Tracking Project. https://covidtracking.com/data, accessed September 2020.

[5] US Centers for Disease Control and Prevention. https://www.cdc.gov/coronavirus/2019-ncov/need-extra-precautions/people-with-medical-conditions.html, accessed September 2020.

[6] US Centers for Disease Control and Prevention. https://www.cdc.gov/nchs/fastats/flu.html, accessed September 2020.

[7] World Health Organization. https://www.who.int/health-topics/coronavirus#tab=tab_3, accessed September 2020.

[8] V. Chankong and Y. Y. Haimes. Multiobjective Decision Making: Theory and Methodology. Courier Dover, 2008.

[9] Mohammad Reza Davahli, Waldemar Karwowski, Sevil Sonmez, and Yorghos Apostolopoulos. The hospitality industry in the face of the covid-19 pandemic: Current topics and research methods. International Journal of Environmental Research and Public Health, 17(20), 2020.

[10] Michael Dellnitz, Oliver Schütze, and Thorsten Hestermeyer. Covering pareto sets by multilevel subdivision techniques. Journal of Optimization, Theory and Applications, 124(1), 2005.

[11] Luca Dell’Anna. Solvable delay model for epidemic spreading: the case of covid-19 in italy. Scientific Reports, 10, 2020.

[12] Ali Eshragh, Saed Alizamir, Peter Howley, and Elizabeth Stojanovski. Modeling the dynamics of the covid-19 population in australia: A probabilistic analysis. PLOS ONE, 15(10):1–19, 10 2020.

[13] Brauer F. Some simple epidemic models. Math Biosci Eng, 3:1, 2006.

[14] Lin F, Muthuraman K, and Lawley M. An optimal control theory approach to non-pharmaceutical interventions. BMC Infect Dis, 10:32, 2010.

[15] Bjoern Goldenbogen, Stephan O. Adler, Oliver Bodeit, Judith AH Wodke, Aviv Korman, Lasse Bonn, Ximena Martinez de la Escalera, Johanna E L Haffner, Maria Krantz, Maxim Karnetzki, Ivo Maintz, Lisa Mallis, Rafael U Moran Torres, Hannah Prawitz, Patrick Segelitz, Martin Seeger, Rune Linding, and Edda Klipp. Geospatial precision simulations of community confined human interactions during sars-cov-2 transmission reveals bimodal intervention outcomes. medRxiv, 2020.

[16] Sha He, Sanyi Tang, and Libin Rong. A discrete stochastic model of the covid-19 outbreak: Forecast and control. Mathematical Biosciences and Engineering, 17:2792–2804, 01 2020.

[17] Luzie Helfmann, Natasa Djurdjevac Conrad, Ana Djurdjevac, Stefanie Winkelmann, and Christof Schütte. From interacting agents to density-based modeling with stochastic pdes. to appear in Communications in Applied Mathematics and Computational Science, 2020.

[18] Klaus-Dieter Hungenberg and Michael Wulkow. Modeling and Simulation in Polymer Reaction Engineering: A Modular Approach. Wiley VCH Verlag, 2018.

[19] Markus Kantner and Thomas Koprucki. Beyond just “flattening the curve”: Optimal control of epidemics with purely non-pharmaceutical interventions. 2004.09471v3, 2020.

[20] H. W. Kuhn and A. W. Tucker. Nonlinear programming. In Proceedings of the Second Berkeley Symposium on Mathematical Statistics and Probability, pages 481–492, Berkeley, Calif., 1951. University of California Press.

[21] M Linden, S Mohr, J Dehing, J Mohring, M Meyer-Hermann, I Pigeot, A Schöbel, and V Priesemann. ü berschreitung der Kontaktnachverfolgungskapazität gefährdet die Eindämmung von covid-19. November 2020.

[22] Chinwendu Madubueze, Dachollom Sambo, and Isaac Onwubuya. Controlling the spread of covid-19: Optimal control analysis. 06 2020.

[23] Kaisa Miettinen. Nonlinear Multiobjective Optimization. Springer, 1999.

[24] Sebastian A Müller, Michael Balmer, Billy Charlton, Ricardo Ewert, Andreas Neumann, Christian Rakow, Tilmann Schlenther, and Kai Nagel. Using mobile phone data for epidemiological simulations of lockdowns: government interventions, behavioral changes, and resulting changes of reinfections. medRxiv, doi:10.1101/2020.07.22.20160093, 2020.

[25] Sebastian A. Müller, Michael Balmer, William Charlton, Ricardo Ewert, Andreas Neumann, Christian Rakow, Tilmann Schlenther, and Kai Nagel. A realistic agent-based simulation model for covid-19 based on a traffic simulation and mobile phone data. 2011.11453, 2020.

[26] Shah N, Suthar A, and Jayswal E. Control strategies to curtail transmission of covid-19. medRxiv. https://doi.org/10.1101/2020.04.04.20053173, 2020.

[27] Philip Nadler, Shuo Wang, Rossella Arcucci, Xian Yang, and Yike Guo. An epidemiological modelling approach for covid-19 via data assimilation. European Journal of Epidemiology, 35(8), 2020.

[28] Sebastian Peitz and Michael Dellnitz. A survey of recent trends in multiobjective optimal control—surrogate models, feedback control and objective reduction. Mathematical and Computational Applications, 23(2):30, Jun 2018.

[29] T.A. Perkins and G. Espana. Optimal control of the covid-19 pandemic with non-pharmaceutical interventions. Bull Math Biol, 82, 118, 2020.

[30] Robert Koch-Institut. COVID-19-Dashboard. www.corona.rki.de, 2020. Last accessed: 2020-11-22.

[31] Susanna Roeblitz and Peter Deuflhard. A Guide to Numerical Modelling in Systems Biology. Springer, 2015.

[32] Oliver Schütze, Katrin Witting, Sina Ober-Blöbaum, and Michael Dellnitz. Set oriented methods for the numerical treatment of multiobjective optimization problems. In Emilia Tantar, Alexandru-Adrian Tantar, Pascal Bouvry, Pierre Del Moral, Pierrick Legrand, Carlos A. Coello Coello, and Oliver Schütze, editors, EVOLVE-A Bridge between Probability, Set Oriented Numerics and Evolutionary Computation, pages 187–219. Springer, Berlin Heidelberg, Berlin, Heidelberg, 2013.

[33] Senozon. Mobility Pattern Recognition (MPR) und Anonymisierung von Mobilfunk-daten. https://senozon.com/wp-content/uploads/Whitepaper_MPR_Senozon_DE.pdf, 2020. accessed: 2020-7-21.

[34] Timo Smieszek. A mechanistic model of infection: why duration and intensity of contacts should be included in models of disease spread. Theors. Biol. Med. Model., 6:25, November 2009.

[35] Timo Smieszek. Models of epidemics: how contact characteristics shape the spread of infectious diseases. PhD thesis, Ph.D. thesis, ETH Zurich, Switzerland, 2010.

[36] US Centers for Disease Control and Prevention. https://www.cdc.gov/coronavirus/2019-ncov/covid-data/investigations-discovery/hospitalization-death-by-age.html, accessed September 2020.

[37] Stefanie Winkelmann and Christof Schütte. Stochastic Dynamics in Computational Biology. Frontiers in Applied Dynamical Systems (accepted for publication). Springer, 2020.

[38] World Health Organization. WHO numbers. https://covid19.who.int/, accessed September 2020.

[39] Michael Wulkow. Computer aided modeling of polymer reaction engineering—the status of predici, I -Simulation. Macromolecular Reaction Engineering, 2:461–494, 11 2008.

[40] A. Yousefpour, H. Jahanshahi, and A. Bekiros. Optimal policies for control of the novel coronavirus disease (covid-19) outbreak. Chaos Solitons Fractals, 136:109883, 2020.

[41] Zhang Z, Zeb A, Hussain S, and Alzahrani E. Dynamics of covid-19 mathematical model with stochastic perturbation. Adv Differ Equ., 1, 2020.

